# The smartphone: an evolution or revolution in virtual patient healthcare during and beyond the COVID-19 pandemic ? An evaluation and comparison of the smartphone against other currently available wearable technologies in a secondary care setting during the COVID-19 pandemic

**DOI:** 10.1101/2020.11.06.20223206

**Authors:** Asif Raza, Subhabrata Mukherjee, Vikash Patel, Naila Kamal, Ewa Lichtarowicz-Krynska

## Abstract

Smartphones are now commonly used, for virtual outpatient consultations, to help reduce disease transmission during the COVID-19 pandemic. Nosocomial spread of COVID 19 and hospital acquired infections are usually from staff or students to patients. Reducing non-essential staff numbers on ward rounds may reduce the risk.

We describe the novel use of smartphones, with Microsoft Teams, to live stream inpatient interactions, radiological images, pathology results, charts and patient review between an office-based and ward team (virtual ward round) and for teaching medical students in secondary care. After Research and Ethics, Digital services and Information Governance approval we compared a smartphone and head-worn device (Realwear HMT-1). Data collection was by participant questionnaire. Statistical analysis was performed using the Mann – Whitney test.

There was no statistically significant difference in audio and video feed quality between the smart phone (p value = 0.3) and Realwear device (p value = 0.41). However the smartphone was preferred during ward rounds and was 85% cheaper than the Realwear device. Urology medical staff numbers on the ward were reduced by 50%. Ward round efficiency improved as administrative tasks could be performed by the office team during the virtual ward round.

Virtual ward rounds using smartphones can facilitate remote communication between staff, students and patients. Staff in isolation or shielding can also assist front line colleagues from home. Smarter use of the smart phone may help reduce staff numbers on wards and reduce the number COVID-19 and nosocomial infections, potentially reducing morbidity and mortality locally and globally.

## Introduction

There has been an exponential increase in telemedicine, [1,2] mobile health platforms (mhealth) [3-5] and virtual clinics during the COVID-19 pandemic.[6] Up to 87% of doctors use their smartphone for work related tasks.[7]

The necessity for certain patients or staff to shield or isolate during the first wave of COVID-19 e.g. higher risk Black, Asian and Minority Ethnic (BAME) staff, highlighted limitations in communication platforms between staff, and between doctors and patients. This could have contributed to higher patient morbidity and mortality particularly in care homes.[8] Medical student teaching suffered as hospital access was restricted during the height of the pandemic. Newer but often costly technologies are playing an increasing role in medical education.[9] However concerns remain about smart devices and data privacy and security, network and Wi-Fi capabilities, costs, user acceptance and bacterial contamination. The Information Commissioner’s Office (UK) enforce General Data Protection Regulations (GDPR) [10] and NHS Digital has approved Microsoft teams (MS Teams), a secure platform, with 256-bit Advanced Encryption Standard (AES) protection of data for routine use by clinicians. Staff smartphones may be colonised with MRSA and VRE.[11-13] However smartphone and tablet touchscreens are associated with lower infection levels than standard touchpad keyboards in hospitals.[14] Decontamination methods include antibacterial screens (silver coated), alcohol-based wipes (70% isopropyl alcohol), Clinell wipes and Ultraviolet light(UVC). [15-17]

Performing virtual ward rounds may help reduce nosocomial infection risk including COVID 19 transmission. Up to 1.4 million patients are affected by non-COVID-19 hospital acquired nosocomial infections annually increasing morbidity and mortality particularly in the elderly and frail. [18] Up to 20%-70% patient files and charts harbour infection due to frequent handling by staff [19-21] COVID-19 spreads by droplet transmission, poor hand hygiene and contact between staff and patients [22] and up to 80% of staff may be asymptomatic carriers of COVID-19 [23] risking nosocomial spread from staff to patients. [24 – 27]

Virtual ward rounds have been reported previously, with less staff on the ward, using a double telepresence robot (iPad on a trolley),[28] Microsoft Hololens [29] and laptops carried onto the ward.[30] The adoption of an office based team (OBT) and remote ward team (RWT) in our practice reduced staff numbers by 50% on the ward round.

We wished to determine the most practical, user friendly and cost-effective wearable device that would allow staff to safely communicate remotely during virtual ward rounds, assess device practicality, use in theatres by surgeons, and for remote teaching of medical students during and after COVID 19.

## Method

A google search revealed several commercially available wearable devices that live stream video feed. Devices required MS Teams functionality (without using third party apps for connection to MS teams) to enable live video streaming and two-way audio communication. Selected devices were tested in mock clinical scenarios (Paediatric Resuscitation and Sim-Man simulation training) and subsequently tested with patients. Fig 1

**FIG 1.**
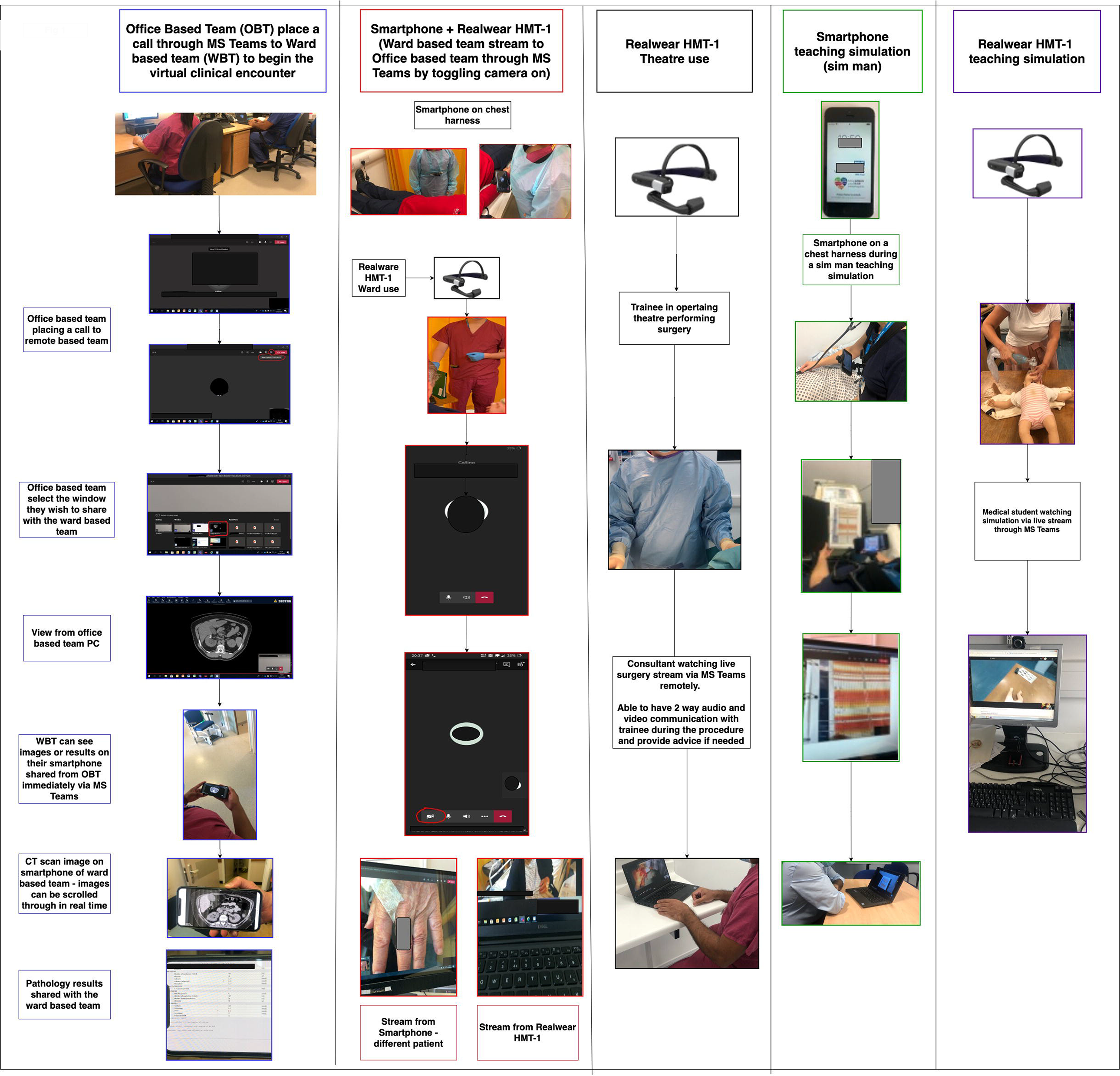
survey, summary of results of smart devices evaluated.

Our local Research and Ethics committee approved the study as a service evaluation and proof of concept study. The local Information Governance Officer (IGO) and local Digital services team approved live streaming via MS Teams. No recording or storing of video or images during or after the mock or real-life patient interaction was allowed for the study unless separate written consent was obtained for video or images to be kept for teaching purposes or publication. Informed consent was taken from patients, students and staff prior to streaming.

Inclusion criteria included all genders, over 18 years old, all socio-economic groups and medical or surgical condition not requiring emergency treatment. Exclusion criteria included patients unable to give valid informed consent due to a language barrier or incapacity, patients with life threatening emergencies, or those who were septic or confused on admission.

An online questionnaire was completed by the wearer of the device (RWT) and receivers of the live video feed stream (OBT, students) after streaming the interaction. (Table 1). Likert scoring was used to assess wearer and receiver satisfaction with the devices and streaming quality. Patients were asked to complete a post streaming satisfaction questionnaire.

**Table 1.**
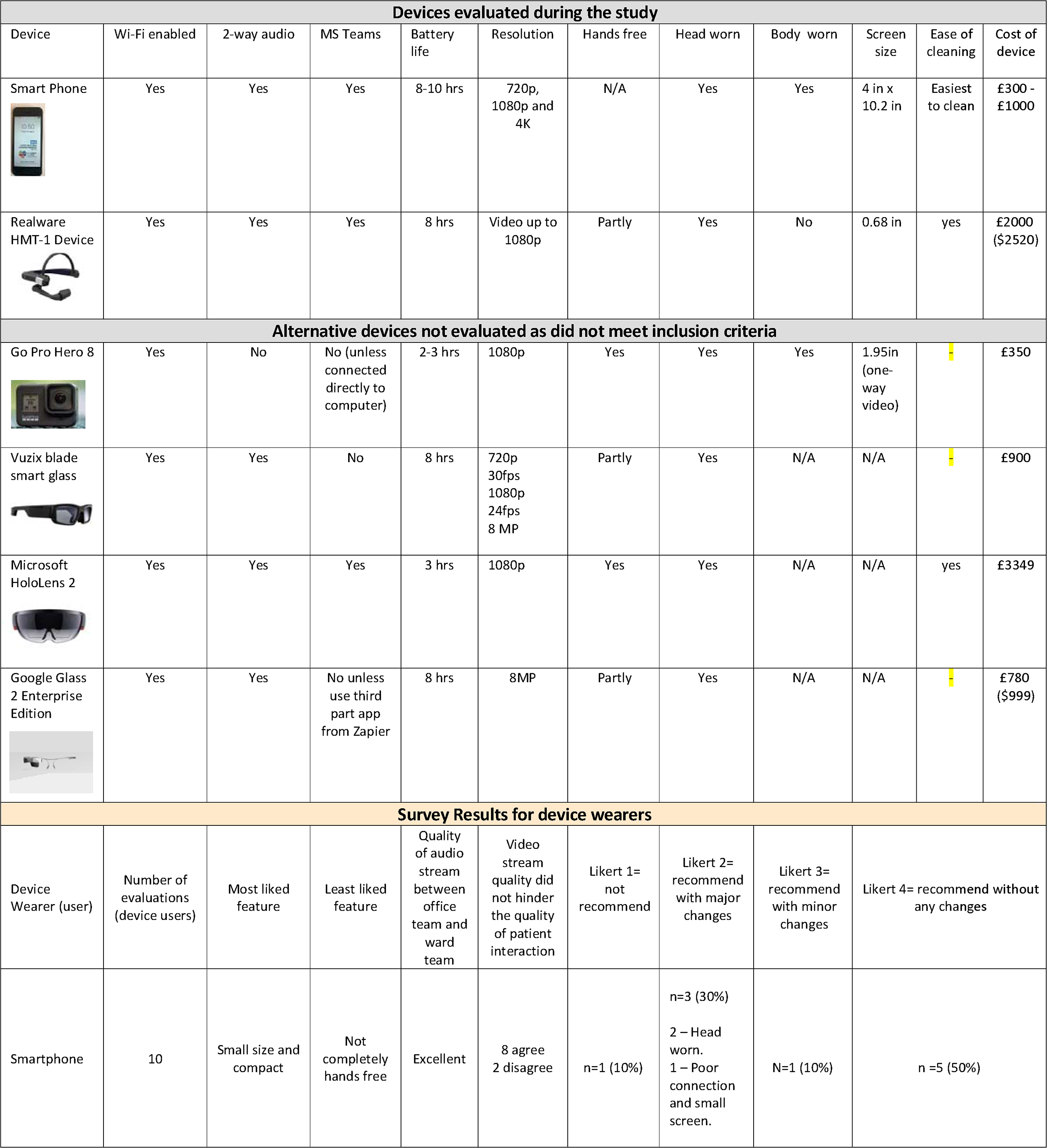
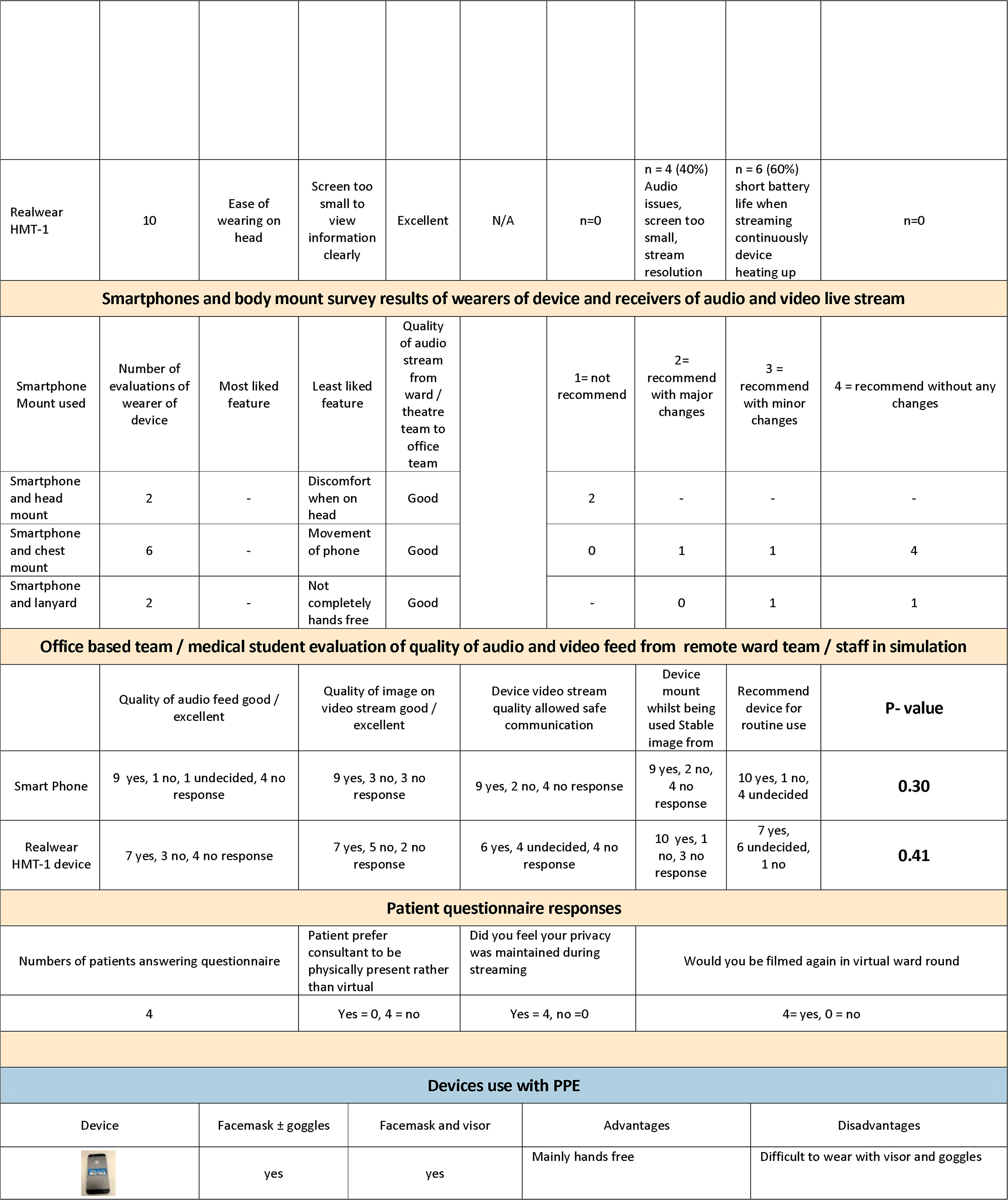
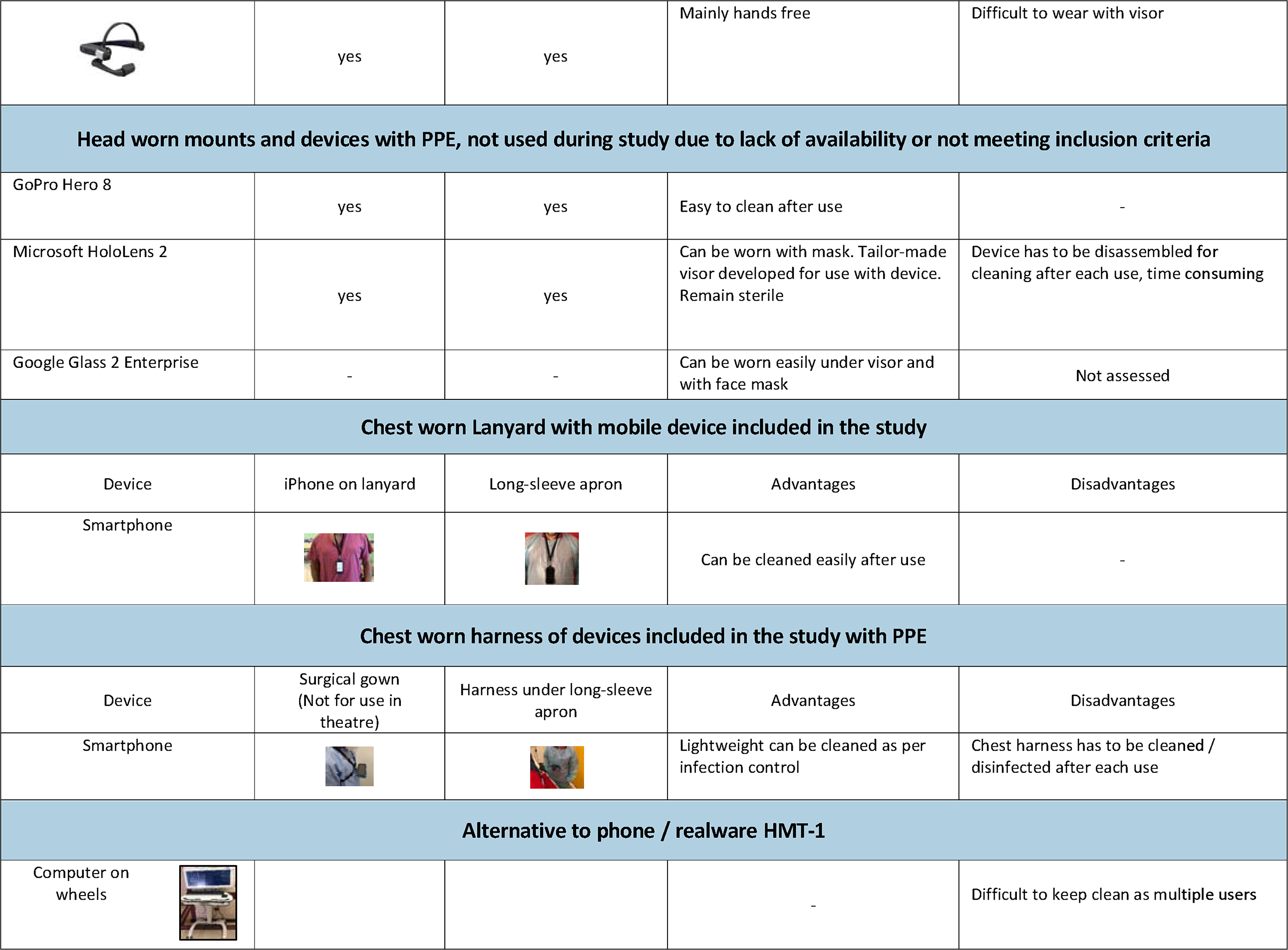
Device survey, summary of results of smart devices evaluated for virtual patient interaction and teaching simulations, PPE and devices

| Devices evaluated during the study |  |  |  |  |  |  |  |  |  |  |  |
| --- | --- | --- | --- | --- | --- | --- | --- | --- | --- | --- | --- |
| Device | Wi-Fi enabled | 2-way audio | MS Teams | Battery life | Resolution | Hands free | Head worn | Body worn | Screen size | Ease of cleaning | Cost of device |
| Smart Phone | Yes | Yes | Yes | 8-10 hrs | 720p, 1080p and 4K | N/A | Yes | Yes | 4 in x 10.2 in | Easiest to clean | £300 - £1000 |
| Realware HMT-1 Device | Yes | Yes | Yes | 8 hrs | Video up to 1080p | Partly | Yes | No | 0.68 in | yes | £2000 (\$2520) |
| Alternative devices not evaluated as did not meet inclusion criteria |  |  |  |  |  |  |  |  |  |  |  |
| Go Pro Hero 8 | Yes | No | No (unless connected directly to computer) | 2-3 hrs | 1080p | Yes | Yes | Yes | 1.95in (one-way video) |  | £350 |
| Vuzix blade smart glass | Yes | Yes | No | 8 hrs | 720p 30fps<br>1080p 24fps<br>8 MP | Partly | Yes | N/A | N/A |  | £900 |
| Microsoft HoloLens 2 | Yes | Yes | Yes | 3 hrs | 1080p | Yes | Yes | N/A | N/A | yes | £3349 |
| Google Glass 2 Enterprise Edition | Yes | Yes | No unless use third part app from Zapier | 8 hrs | 8MP | Partly | Yes | N/A | N/A | | £780 (\$999) |
| Survey Results for device wearers |  |  |  |  |  |  |  |  |  |  |  |
| Device Wearer (user) | Number of evaluations (device users) | Most liked feature | Least liked feature | Quality of audio stream between office team and ward team | Video stream quality did not hinder the quality of patient interaction | Likert 1= not recommend | Likert 2= recommend with major changes | Likert 3= recommend with minor changes | Likert 4= recommend without any changes |  |  |
| Smartphone | 10 | Small size and compact | Not completely hands free | Excellent | 8 agree<br>2 disagree | n=1 (10%) | n=3 (30%)<br>2 – Head worn.<br>1 – Poor connection and small screen. | N=1 (10%) | n =5 (50%) |  |  |

|  |  |  |  |  |  |  |  |  |  |
| --- | --- | --- | --- | --- | --- | --- | --- | --- | --- |
| Realwear HMT-1 | 10 | Ease of wearing on head | Screen too small to view information clearly | Excellent | N/A | n=0 | n = 4 (40%) Audio issues, screen too small, stream resolution | n = 6 (60%) short battery life when streaming continuously device heating up | n=0 |

Smartphones and body mount survey results of wearers of device and receivers of audio and video live stream
| Smartphone Mount used | Number of evaluations of wearer of device | Most liked feature | Least liked feature | Quality of audio stream from ward / theatre team to office team | 1= not recommend | 2= recommend with major changes | 3 = recommend with minor changes | 4 = recommend without any changes |
| --- | --- | --- | --- | --- | --- | --- | --- | --- |
| Smartphone and head mount | 2 | - | Discomfort when on head | Good | 2 | - | - | - |
| Smartphone and chest mount | 6 | - | Movement of phone | Good | 0 | 1 | 1 | 4 |
| Smartphone and lanyard | 2 | - | Not completely hands free | Good | - | 0 | 1 | 1 |

Office based team / medical student evaluation of quality of audio and video feed from remote ward team / staff in simulation
|  | Quality of audio feed good / excellent | Quality of image on video stream good / excellent | Device video stream quality allowed safe communication | Device mount whilst being used Stable image from | Recommend device for routine use | P- value |
| --- | --- | --- | --- | --- | --- | --- |
| Smart Phone | 9 yes, 1 no, 1 undecided, 4 no response | 9 yes, 3 no, 3 no response | 9 yes, 2 no, 4 no response | 9 yes, 2 no, 4 no response | 10 yes, 1 no, 4 un decided | **0.30** |
| Realwear HMT-1 device | 7 yes, 3 no, 4 no response | 7 yes, 5 no, 2 no response | 6 yes, 4 undecided, 4 no response | 10 yes, 1 no, 3 no response | 7 yes, 6 undecided, 1 no | **0.41** |

Patient questionnaire responses
| Numbers of patients answering questionnaire | Patient prefer consultant to be physically present rather than virtual | Did you feel your privacy was maintained during streaming | Would you be filmed again in virtual ward round |
| --- | --- | --- | --- |
| 4 | Yes = 0, 4 = no | Yes = 4, no = 0 | 4 = yes, 0 = no |

Devices use with PPE
| Device | Facemask ± goggles | Facemask and visor | Advantages | Disadvantages |
| --- | --- | --- | --- | --- |
|  | yes | yes | Mainly hands free | Difficult to wear with visor and goggles |
|  | yes | yes | Mainly hands free | Difficult to wear with visor |
| **Head worn mounts and devices with PPE, not used during study due to lack of availability or not meeting inclusion criteria** |  |  |  |  |
| GoPro Hero 8 | yes | yes | Easy to clean after use | - |
| Microsoft HoloLens 2 | yes | yes | Can be worn with mask. Tailor-made visor developed for use with device. Remain sterile | Device has to be disassembled for cleaning after each use, time consuming |
| Google Glass 2 Enterprise | - | - | Can be worn easily under visor and with face mask | Not assessed |
| **Chest worn Lanyard with mobile device included in the study** |  |  |  |  |
| Device | iPhone on lanyard | Long-sleeve apron | Advantages | Disadvantages |
| Smartphone |  |  | Can be cleaned easily after use | - |
| **Chest worn harness of devices included in the study with PPE** |  |  |  |  |
| Device | Surgical gown (Not for use in theatre) | Harness under long-sleeve apron | Advantages | Disadvantages |
| Smartphone |  |  | Lightweight can be cleaned as per infection control | Chest harness has to be cleaned / disinfected after each use |
| **Alternative to phone / realware HMT-1** |  |  |  |  |
| Computer on wheels |  |  | - | Difficult to keep clean as multiple users |

The (OBT) and students were blinded to the device being used by the (RWT). The device worn was randomly allocated to the (RWT) by using an online randomisation programme (Graph Pads: Quickcalc) The same mock clinical scenario was used for teaching medical students (Fig 1.) for each device to ensure devices could be compared in the same location ensuring Wi-Fi signal strength and network characteristics were similar for both devices.

A sim free hospital approved smartphone was used at the request of our Digital Services department in preference to a personal smartphone. The selected devices were connected to the staff only hospital Wi-Fi, registered to a dedicated network via MAC address and Wi-Fi password was WPA2 encrypted. Device access was password protected.

To ensure privacy of ward patients not in the study bedside curtains were drawn around the patient, in the ward, before live streaming started and undrawn when streaming stopped. The wearer of the device would place the device in standby mode, or switch it off before curtains were withdrawn, and moving to the next patient.

The statistical analyses compared responses for the smartphone to the Realwear device. Due to the ordinal nature of the recommendation measure, the analyses were performed using the Mann-Whitney test. Separate analyses were performed for wearers and receivers.

## Results

Table 1 summary of results : type of device used, questionnaire results, different and use of devices with PPE.

The smartphone was most practical in a ward setting. Most respondents preferred ease of use, the audio and video feed, larger screen, being able to easily see radiological images and results on the smartphone compared to the Realwear device. Most of the negative comments when using a smartphone were due to the harness or mount rather than smartphone.

The Realwear device was preferred for theatre use but not suitable for wards or teaching as the noise cancellation on the device did not allow patients or other staff voices to be picked up by its microphone and therefore heard by the OBT or students. The resolution of the images being viewed remotely were not high enough definition to teach surgeons comprehensively however enough anatomical detail was visible to provide advice to the trainee during a basic surgical procedure.

The results of the online questionnaire suggested that there was no statistically significant difference between the Phone and Realwear for either group of study participants. None of the patients objected to being live streamed or the concept of the virtual ward round.

## Discussion

Limitations in our study included the small number of study participants, the use of older model hospital iphones without a sim, with lower screen resolution, smaller screen size and shorter battery life than current models. Evaluation of the Realwear device with proposed future software updates, with better image resolution and a fix for noise cancellation, would have allowed a better comparison with the smartphone.

The local information governance and Digital services team did not approve video recording or taking pictures on the device. This limited the full evaluation of devices as teaching aids. Our local digital health services team had concerns about the amount of data that would be generated from multiple device use, saving video clips and were and how long the data would be kept i.e. cloud or local servers.

Network coverage was patchy in certain parts of the hospital leading to poor streaming quality and occasional dropped calls. The local trust was conducting a network survey during the study with plans to install multiple wireless access points in areas of poor network coverage.

Our infection control team raised concerns about the disinfection of chest harnesses however these can be worn under sterile gowns however would still need to be cleaned as per local trust guidelines. We used lanyards as an alternative as these are easier to disinfect after use and less bulky to wear.

## Conclusion

The involvement of the local digital services and Information governance teams is essential before implementing virtual streaming of patients and their clinical information. A local agreement at the outset to record as well as live stream would be beneficial for student teaching and medical records.

Further research is needed on how remote viewing of live streams by staff or students off site can be kept secure ensuring no breaches in patient privacy, whether virtual ward rounds will improve ward round efficiency, allow better experiential learning for junior staff performing consultant free ward rounds and procedures, whether the adoption of office and ward-based teams using a smartphone for secure streaming of information in real time can help reduce the risk of nosocomial infection transmission. Further development of more infection friendly harnesses for use with smart phones needs to be developed.

Smarter use of the smartphone during and after COVID -19 pandemic could save not only overall costs for smart technologies but also reduce mortality and morbidity.

However for the benefits to be realised clinician engagement of a new model of remote based management of patients and teaching of students is essential.

## Data Availability

Anonymised Data will be made available on request

## Acknowledgements

I would like to thank Mr Conrad Jaggwe Technical Lead and Facilities officer, Postgraduate Medical Education Dept, London Northwest Healthcare University NHS Trust for his technical assistance and advice on how to set up these devices, the Digital services team for providing the smartphones for evaluation, the Education fellows, Urology trainees and Medical students who assisted in testing and assessing devices during mock and real-life patient interactions. I would also like to thank the Realwear company for providing a device for us to evaluate during the study.

